# Designing a Perioperative Mind-Body Intervention for Peripheral Vascular Interventions

**DOI:** 10.1101/2024.06.07.24308071

**Authors:** C. Y. Maximilian, Darshan Mehta, Anahita Dua, Antonia Stephen, Alex M. Bruce, Aynsley Forsythe, Hovig V. Chitilian, Erik J. Bringle, James C. Simpson, Katherine M. Parady, Lisa A. McNeil, Margaret A. Baim, Matthew J. Eagleton, David C. Chang, Gloria Y. Yeh

## Abstract

**Background:** Peripheral vascular interventions (PVIs) performed under procedural sedation and analgesia (PSA) can be associated with anxiety and poor compliance with patient instructions during surgery. Mind-body interventions (MBIs) such as meditation have demonstrated the potential to decrease perioperative anxiety, though this area is understudied, and no tailored interventions have been developed for the vascular surgical patient population.

**Objectives:** We aimed to design a perioperative MBI that specifically targeted vascular surgical patients undergoing PVIs under PSA. We sought to perform this in a scientifically rigorous, multi-disciplinary collaborative manner.

**Methods:** Following the Obesity-Related Behavioral Intervention Trials (ORBIT) model, we designed (Phase 1a) and then refined (Phase 1b) a MBI for patients undergoing PVIs under PSA to decrease perioperative anxiety and sedation and facilitate patient intraoperative compliance. Phase 1a involved a literature review, informal information gathering and synthesis, and drafting a preliminary protocol for a perioperative MBI. Phase 1b involved assembling a multi-disciplinary expert panel of perioperative and mind-body clinicians and researchers to improve the MBI using an iterative, modified Delphi approach.

**Results:** The modified Delphi process was completed, and a consensus was reached after three iterations. The resulting MBI consisted of two seven-minute preoperative guided meditations on the day of surgery, including diaphragmatic breathing, body scans, and guided imagery emphasizing awareness of the ipsilateral leg where the vascular surgery was performed. A document delineating the integration of the MBI into the operating room workflow was produced, including details regarding the intervention’s timing, duration, and modality.

**Conclusion:** Using a multi-specialty expert panel, we designed a novel MBI in the form of a guided meditation with elements of mindfulness and guided imagery to decrease anxiety and increase intraoperative compliance for patients undergoing PVIs under PSA. A prospective pilot study is being planned to test the program’s feasibility.

## Introduction

Perioperative anxiety is a state of anxiety that patients often experience before and during their surgical procedure related to anesthesia or surgery, including fears about their medical condition, adverse events, or complications.^1,2^ This anxiety is extremely common, even in individuals without any prior diagnosis of anxiety disorder, and has been associated with perioperative hemodynamic instability and, correspondingly, worse procedural outcomes.^1,3^ Perioperative anxiety can be further compounded for patients with an anesthetic plan of procedural sedation and analgesia (PSA, also known as conscious sedation) as opposed to general anesthesia because they remain continuously aware of perioperative activity.^4,5^ For patients undergoing peripheral vascular interventions (PVIs), PSA is often the preferred standard-of-care, because it avoids the cardiopulmonary stress of intubation, though patients need to be awake enough to comply with essential perioperative instructions from proceduralists such as holding the ipsilateral leg still and breath holds at certain intervals.^6^ Perioperative anxiety can be especially problematic in those undergoing PVI’s since it can impact patients’ ability to comply with these intra-operative procedural instructions, which can result in repeated imaging attempts and thus increased intravenous contrast administration and radiation exposure to both the patients and the operative room providers. For such patients, the pharmacological balance of anesthesia between patient comfort/anxiety reduction and intraoperative compliance can be challenging, as oversedation can not only result in cardiopulmonary comprise but also involuntary disinhibition, which can prompt intraoperative conversion to general anesthesia or even on-table abortion of the procedure.^7^

Mind-body interventions (MBIs) have been shown to have anxiolytic and resulting positive effects on hemodynamic parameters in both the inpatient and outpatient settings.^8,9^ Prior perioperative MBIs have ranged from relaxation techniques and music therapy to hypnosis, with mixed but generally favorable results in terms of postoperative pain, anxiety, and even hemodynamics such as modest decreases in heart rate and diastolic blood pressure.^10-17^ Additionally, a study comparing pharmacological sedation to pharmacological sedation with adjunctive hypnosis showed that there were potential cost savings even if the group with adjunctive hypnosis consumed more operating room time.^18^ In a ninety-patient trial of patients undergoing spinal fusion surgery, perioperative cognitive behavioral therapy (CBT) was also shown to result in improved pain tolerance and improved postoperative mobility.^19^ More recently, preoperative meditation has demonstrated encouraging results; three sessions of preoperative pre-prandial Rajyoga meditation in cardiac surgery patients not only resulted in less postoperative anxiety but also significantly decreased serum cortisol levels compared to a control group.^20^ Other studies in orthopedic surgery have associated similar mind-body interventions with better sleep quality and pain control;^21^ and several trials are still ongoing.^22^

Despite this supportive literature, this area remains understudied. There are several notable challenges to implementing such interventions in the perioperative arena, including time constraints, concomitant periprocedural tasks from various operating room teams, the potential for additional staff to be needed, and the need for more high-quality evidence to support such novel interventions.^23-25^ Furthermore, while prior MBIs trialed perioperatively have shown preliminary promise, to our knowledge, interventions have not been designed for or tailored to a specific surgery. There has yet to be an intervention with content for vascular surgical patients.^17^ Given the intraoperative instructions for patients undergoing PVIs that are unique to vascular surgery, a guided meditation program focused on breath awareness and body scan techniques could potentially synergize with adequate analgesia and sedation goals.^7^ As such, we sought to design a novel perioperative MBI with the goal to decrease anxiety and increase intraoperative compliance for patients undergoing PVIs under PSA.

## Methods

To develop our intervention, we used the framework from the ORBIT model, which is known as a flexible and progressive approach for the creation of novel behavioral treatments.^26^ Given the multitude of stakeholders in the perioperative space, we sought to have input from perioperative nurses, radiation technologists, anesthesiologists, surgeons, integrative medicine clinicians, and researchers, and also vascular surgical patients themselves, to design a mind-body intervention using a multi-disciplinary expert panel with both high fidelity and high likelihood of successful integration into the perioperative space. This paper describes the equivalent of Phase Ia (Design) and Phase Ib (Refine) from the ORBIT model to develop our intervention. This process was deemed exempt by the Mass General Brigham Institutional Review Board.

### Phase Ia (Design)

We began the design of our protocol with a literature review of prior mind-body interventions and anxiolytic therapies used in the perioperative care of surgical patients. We additionally gathered informal information through contact with thought-leaders in the mind-body clinical and research field (through the personal network of our study team, including but not limited to those within our institutions) to solicit feedback. Leaders included the clinical and research directors and researchers at the Benson-Henry Institute for Mind-Body Medicine at Massachusetts General Hospital (MGH), the Osher Center for Integrative Health at Harvard Medical School and Brigham & Women’s Hospital, and leaders in the Mindfulness-Based Stress Reduction program developed at the University of Massachusetts Medical School. In the context of routine clinical care, the lead investigator (CP) had previously sought informal feedback from ten patients who had previously undergone PVIs regarding their perioperative care and how they felt their experience would have been improved.

With the synthesis of this information and support, we modified a preexisting, publicly available breath awareness and body scan guided meditation script by Jon Kabat-Zinn^27^ to tailor it to our target patient population. Given that the typical vascular surgical patient undergoing PVIs is of advanced age and comorbid, we were deliberate about using a trauma-informed perspective when editing our script.^28^ The stated goals of the intervention were to decrease patient anxiety, increase patient compliance with intermittent breath holds, and keep the procedure extremity still.

This early script and the preliminarily proposed logistics of the intervention were presented during two group meetings, each with perioperative staff, including vascular anesthesiologists and nurses, to solicit verbal feedback. Logistical considerations of the MBI included the timing (specifically how to chronologically integrate it into the typical perioperative workflow), duration (the length of the recording(s)). and location (the pre-operative holding area versus in transit to the operating room versus in the operating room). Based on this initial feedback received and with input from the senior study investigators with mind-body research expertise, a proposed protocol for our PVI-specific guided meditation program was developed.

### Phase Ib (Refine)

Expert panel: Formal recruitment for our expert panel was performed through the leadership of the respective departments of integrative medicine, surgery, and anesthesia at MGH to have appropriate representation amongst healthcare providers who care for vascular surgical patients in the perioperative sphere at MGH. We first began via open feedback from the core investigator team (CP, DM, GY) to develop a detailed protocol, which included basic tenets of when, where, and how the meditation program would be implemented in the context of the perioperative encounter. Next, as described below, the input of the larger expert panel was solicited and iteratively incorporated into the design of the proposed MBI. While the program’s stated objectives were to decrease patient anxiety perioperatively and increase patient compliance with on-table instruction intraoperatively, it was emphasized that this intervention should minimize disruption to the existing operating room workflow and avoid adding additional work for the perioperative staff.

We employed a modified Delphi process for the Expert Panel according to the following iterative steps:

1. Documents containing background research, objectives, and the preliminary protocol and preliminary meditation script were distributed to the panel, with highlighted changes from the prior iteration of the program.
2. The panel members reviewed the documents. Then, they provided their assessment of the current version of the intervention via RedCap survey, a secure web-based application that allowed reporting and collating of responses directly to the lead investigator while remaining anonymous to the rest of the panel. This assessment had two parts: numerical ratings on a Likert scale (Figure 1, where 1 corresponded to Strongly Agree and 5 corresponded to Strongly Disagree) and the second was open-ended feedback regarding program elements being debated. These question categories included assessments of potential implications for current perioperative workflow and the potential benefits (to both the patients and healthcare providers) of the proposed intervention.
3. Based on such responses, the lead investigator interviewed individual panel members individually for further elaboration and clarification.
4. The feedback (from steps 2 and 3) was then summarized in a Word document, which included descriptive statistics of the numerical ratings.
5. The core investigator team then convened to review the documents and make appropriate modifications to the program.
6. This cycle was repeated until the expert panel achieved consensus (pre-determined to be a mean score of less than 1.5.

## Results

### Phase 1a (Design)

#### Content of the meditation audio script

Informed by our preexisting clinical knowledge and experience and the informal feedback from our clinical PVI patients, the preexisting guided meditation script was preliminarily modified. Patients mentioned the sensations of pain and cramping most noticeable during obtaining femoral arterial access, but also during balloon angioplasty, and discomfort throughout the procedure when they were asked to keep their ipsilateral leg in the same position for a prolonged period. Based on the synthesis of this information and our literature search, the script was modified to focus on the body scan of the ipsilateral leg (of the procedure) and mindful breath awareness. A focused body scan of the groin area was initially included due to the potential benefit of increased awareness of the site planned for vascular access. After investigator discussion in the context of best trauma-informed perspectives, this component was removed and deemed too emotionally sensitive an area to target. The script was further refined with appropriate language to contextualize the program script as an adaptation of the original and to state the aim of the study to improve perioperative clinical outcomes. We also received recommendations for the voice pacing indicators for the audio recording.

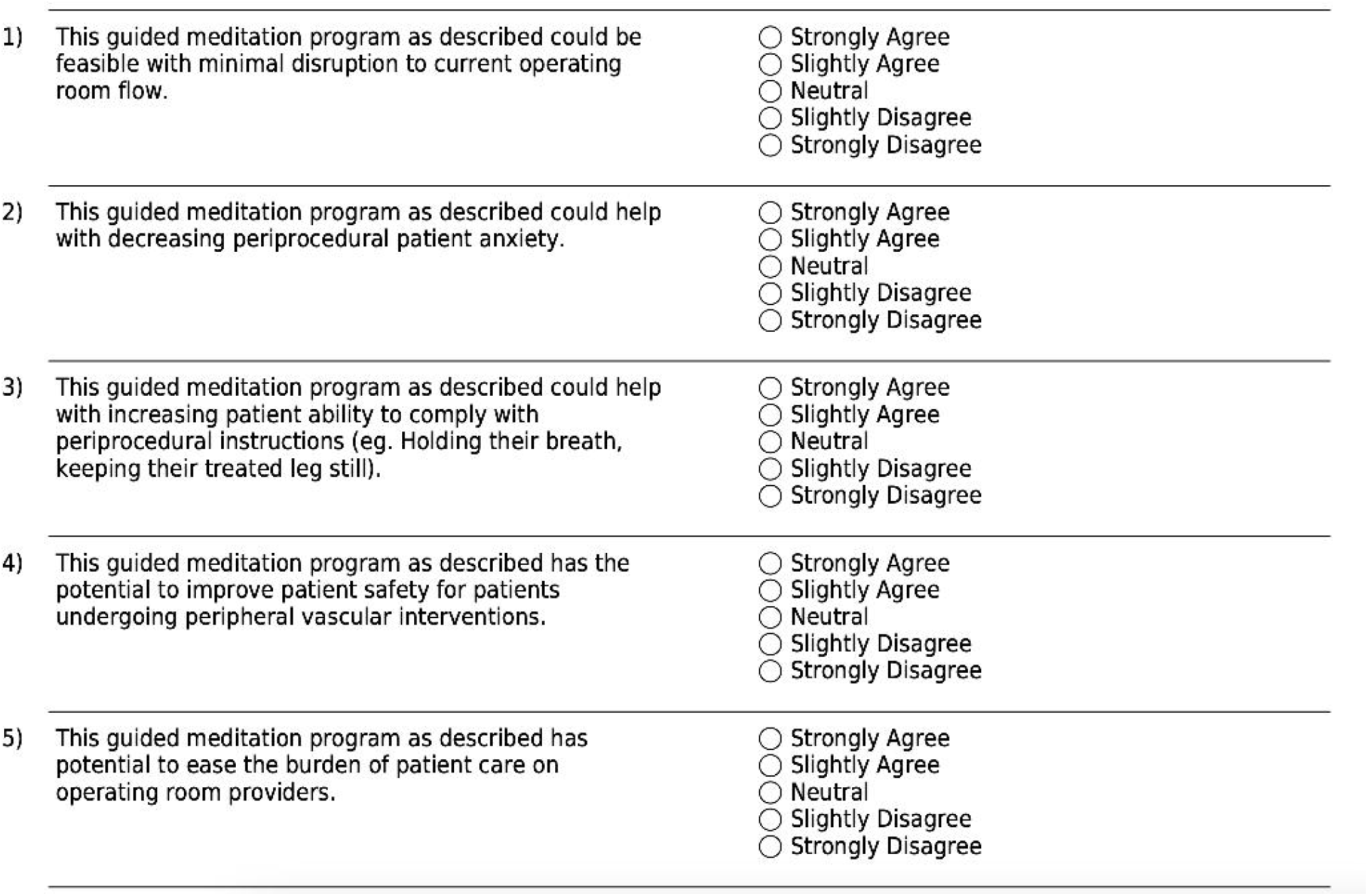

#### Implementation in the perioperative setting

During our meetings with our perioperative colleagues, there were two critical points of deliberation. First was the location of the guided meditation MBI. An advantage of performing the MBI in the preoperative holding area would be more available time, logistically easier implementation, and more flexibility with timing. An advantage to performing the MBI in the operating room is the potential for more durable and temporal benefits of guided meditation due to the decreased lead-in time to the surgical procedure. Having at least a component of the MBI in the preoperative area would take advantage of unused “buffer time” between procedures when the patient is left waiting. Secondly, standardization of anesthesia regimens across PVIs was proposed, which would better allow for inter-PVI comparisons despite the potential need for some anesthesiologists to depart from their usual anesthetic routines. The protocol proposed at this stage remained in the preoperative setting but included the possibility of an intraoperative component. These meetings also confirmed that our expert panel would have appropriate representation from all the key stakeholders.

### Phase 1b (Refine)

#### Expert Panel Recruitment

Twelve members were recruited for the expert panel, comprising two vascular surgeons with experience in clinical trials (AD, ME), one academic endocrine surgeon (AS), two academic vascular anesthesiologists (HC, JS), two vascular operating room nurses (EB, KP), two perioperative care unit nurses (AF, LM), one radiation technologist (AB), an integrative medicine nurse practitioner (MB) and a physician with clinical and research expertise in mind-body medicine (GY). All twelve-panel members were retained throughout the study period.

#### Modified Delphi Panel process

The modified Delphi process was completed, and consensus (pre-determined as a mean score of less than 1.50) was achieved after three iterations. The first round resulted in a mean score of 1.81, the second round in a mean score of 1.55, and the third round in a mean score of 1.45, thus completing the process. (Figure 2) The statement that consistently saw the best score was statement 2: “This guided meditation program as described could help decrease periprocedural patient anxiety.” The statement that saw the most significant improvement between the first and final rounds was statement 5: “This guided meditation program as described has the potential to ease the burden of patient care on operating room providers.” this statement improved from a mean score of 2.0 to 1.5. The cumulative distributions of the panel’s responses are displayed in Figure 3.

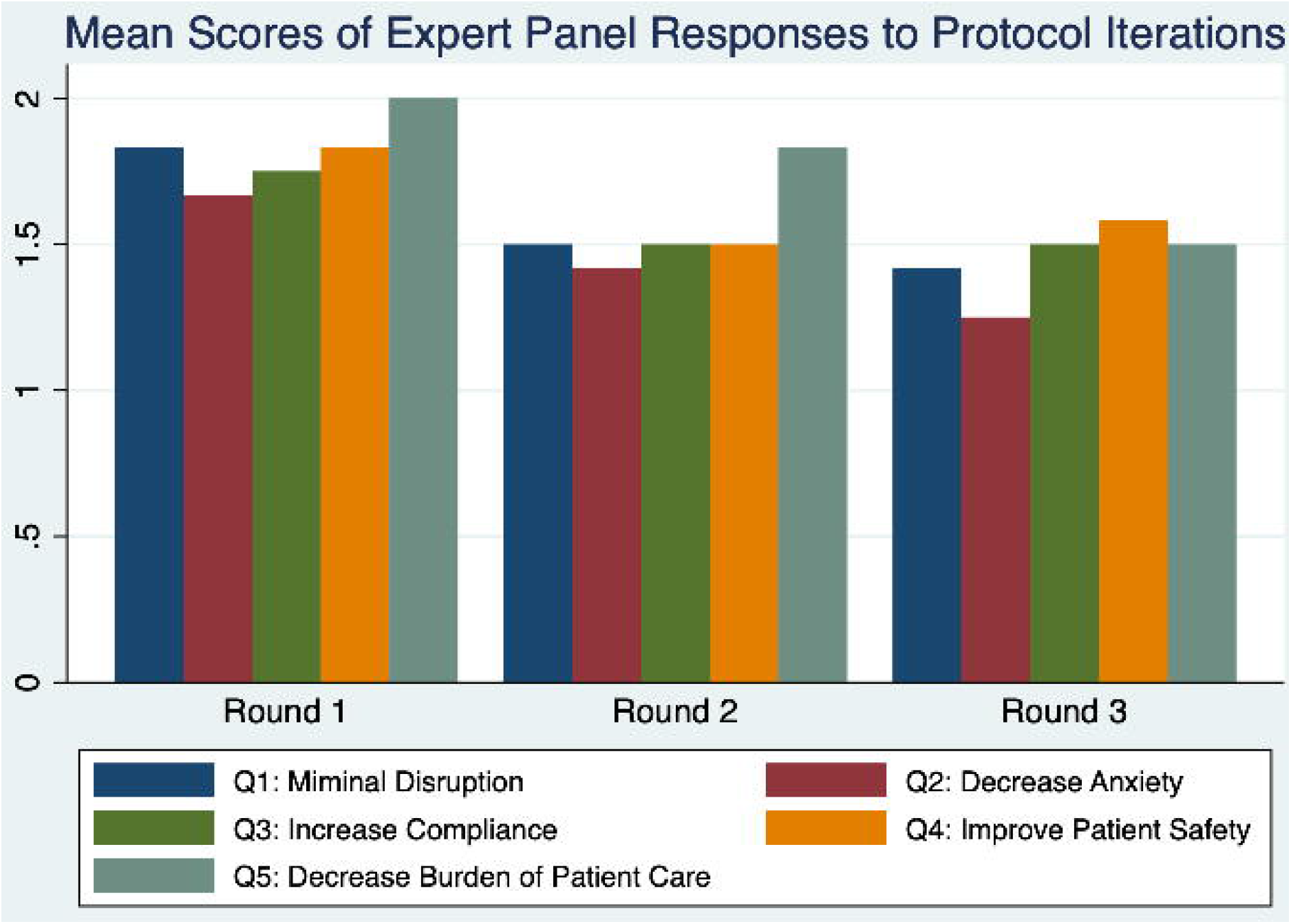

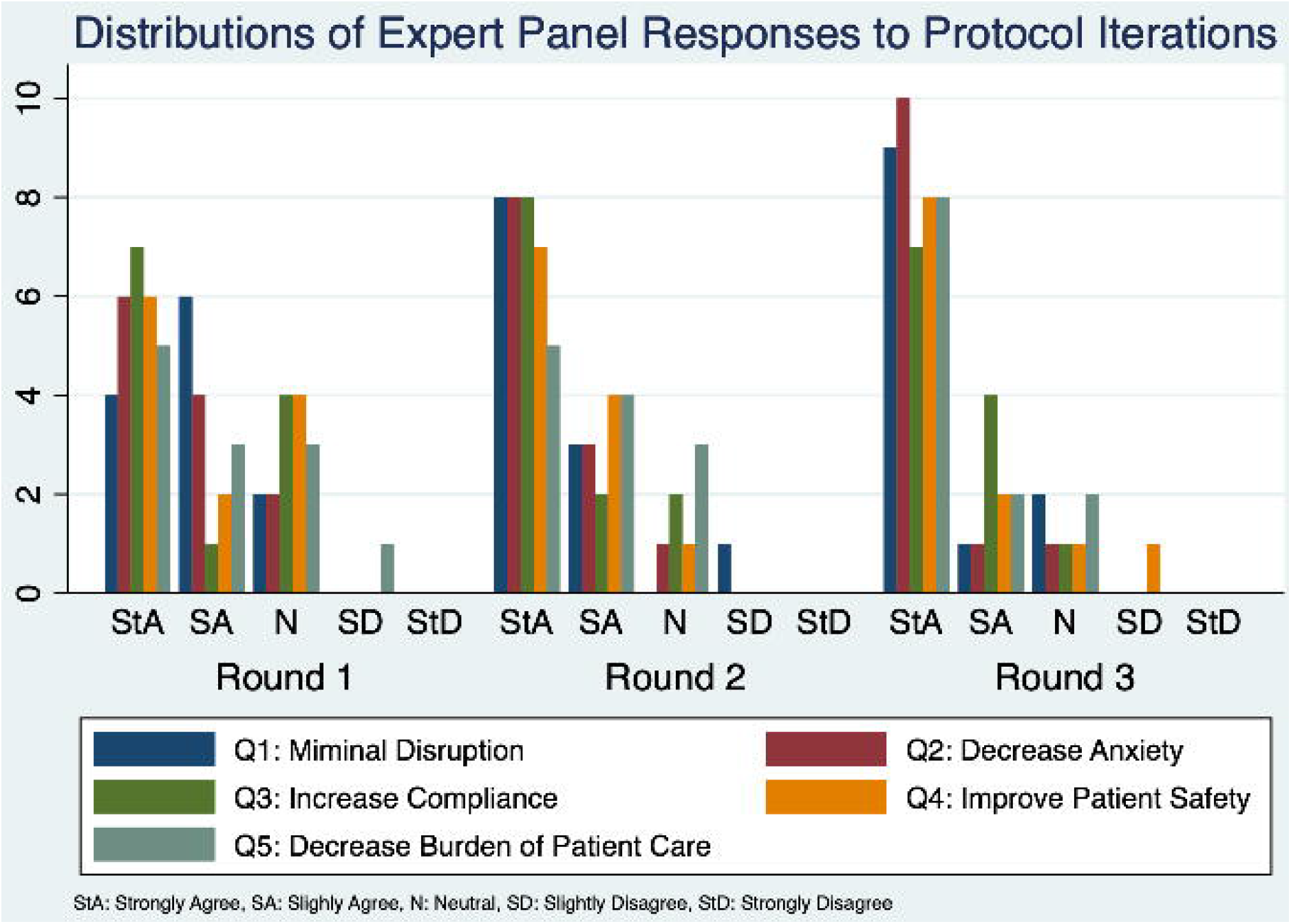

Critical areas of modification/improvement were the timing of existing perioperative workflow, duration of guided meditation sessions, environmental considerations, specific modality, contingencies, script content and distribution across recordings, and recording pace.

#### Final Protocol

The final protocol provided a robust framework for the MBI; below are several fundamental tenets that were refined by the expert panel:

| <u>Protocol Tenet</u> | <u>Preliminary Phase 1a Protocol vs. Modified Phase 1b Protocol</u> | <u>Rationale</u> |
| --- | --- | --- |
| **Perioperative Protocol Implementation** |  |  |
| Timing with existing perioperative workflow | Preliminary: Preoperative, location undefined. | The rationale for a session in the preoperative bay was based on the relative amount of "downtime" for patients in this holding area, which presented a window of opportunity for introducing such intervention without causing any undue delay in care. Furthermore, based on the feedback from the perioperative care unit staff, it was common for patients in the preoperative area to exhibit high levels of anxiety. The second session's rationale for being in the operating room was to reduce the time between the meditation intervention and the |
|  | Modified: Two distinct preoperative sessions: One session in the preoperative holding area and One session after transport to the operating room after patient positioning to be concluded before |  |
|  | surgery | procedure to minimize any potential "washout" and to assist in reorienting the patients to their new environment. |
| Duration of the guided meditation session | Preliminary: Between five and ten minutes | The rationale for seven minutes in the preoperative area was to allow for sufficient time to act as a "primer" at an appropriate pace while at the same time allowing for preoperative clinical care to proceed without delay. The session in the operating room was also seven minutes; it was agreed that this was an appropriate amount of time to overlap with the preoperative activities (including but not limited to the time for the surgeons scrubbing and donning the appropriate surgical gloves and gowns, positioning ultrasound equipment, and performing a preoperative timeout.) |
|  | Modified: Seven minutes for the "primer" session in the preoperative holding area and seven minutes for the operating room session. |  |
| Environment | Preliminary: Provide an environment conducive to guided meditation, otherwise unspecified. | The rationale for dimming and darkening lights and minimizing movement of operating room requirements was to reduce external stimuli that could distract the patient from the guided meditation session. Language that decreased the likelihood of the patients being distracted or even distressed by external stimuli was also inserted into the guided meditation script. |
|  | Modified: Optimize environment for meditation that is appropriate for the perioperative setting by dimming room lights and minimizing staff and equipment movement during the meditation delivery. |  |
| Modality | Preliminary: This will be delivered via headphones, but clinical staff interruptions are allowed if necessary. | The rationale for the MBI to be delivered via a prerecorded script was to increase fidelity and reproducibility compared to delivery from a live practitioner, even if a script was used. Furthermore, open-ear headphones would allow for rapid interruptions if clinically indicated. The tablet device would serve as a real-time monitor for how far along the patient was in the guided meditation, which would be particularly relevant in the case of interruptions. |
|  | Modified: To be explicitly delivered via open-ear headphones with the corresponding script continuously |  |
|  | scrolling on a tablet. |  |
| Contingencies | Preliminary: To have protocols to de-escalate patients in the case of increased anxiety. | The rationale for having contingency protocols is that heightened anxiety in the perioperative setting is prevalent and the target of our intervention. Furthermore, paradoxically increased anxiety has been described during the practice of relaxation techniques (particularly in patients with preexisting anxiety or depressive disorders). However, this is uncommon and has not been described perioperatively.<sup>29</sup> Grounding techniques (which include patients grounding their sense of sight by opening their eyes and their sense of touch by grasping a plush toy within reach) were added to the protocol if needed as a first-line response with pharmacological agents as a second-line therapy. |
|  | Modified: To use grounding of the patients' senses as a de-escalation strategy, in addition to having no restriction on anesthetic practices. |  |
| MBI Content |  |  |
| Script Content | Preliminary: Main script components of diaphragmatic breathing, body scan meditation of ipsilateral leg, and elements of guided imagery<br>Modified: No change | The expert panel did not have further recommendations regarding elements of the script and agreed that the content could be beneficial for the stated goals. Breathing was used as a mindful awareness anchor and attention to the breath, which is later manipulated intraoperatively (breath holds during the procedure). Body scan of ipsilateral leg from toes to thigh focused on the surgical extremity. Guided imagery language envisioning the blood vessels as a river and the surrounding muscles and bones as the riverbank were used to increase accessibility and facilitate meditation of the leg. |
| Distribution of script content | Preliminary: All meditation script content in a single continuous recording<br>Modified: Two separate recordings to deliver appropriate content as a separate primer session and operating room session | Each recording included contextualizing language to reflect the actual location of the patient, as well as encouragement to focus on the meditation regardless of potential surrounding distractors. The first session contained a brief introduction, including the program's purpose, then focused on essential elements of breath awareness, body scan meditation of the ipsilateral leg, and then return to diaphragmatic breathing. The rationale was to provide a primer to the practice before the operating room. Once in the operating room, the |
|  |  | second session started with the same breath awareness and body scan mediation of the ipsilateral leg that had already been introduced. The session also included guided imagery. |
| Pace of recording | Preliminary: Audio script recording to fit within the specified duration<br>Modified: Slow down the audio tempo in certain sections to provide pauses between instructions. | Rationale: To allow time for patients to absorb instruction at the moment and practice accordingly. |

## Discussion

In a systematic approach, we successfully designed a novel MBI in the form of a guided meditation with elements of mindfulness and guided imagery for patients undergoing PVIs under PSA. To our knowledge, this is the first MBI with tailored content specific to a surgical procedure, both within and outside vascular surgery. Additionally, it is the first MBI to specifically target the intra-operative needs of the patient and clinical team during surgery by aiming to increase patient compliance with on-table instruction regarding breath holds and maintaining ipsilateral leg stability.

Our recent nationwide survey of vascular surgeons shows that patient cooperation during PVIs is essential and provides strong rationale for this MBI intervention to have immense benefits regarding perioperative anxiolysis and patient compliance, with few potential risks, particularly compared to pharmacologic agents.^7,30^ Via the established ORBIT framework and the modified Delphi technique, the design of this intervention has resulted in a protocol that has taken into account the complex perioperative setting, time and efficiency of the operating room flow, and the needs and preferences of the perioperative staff and the greatest chance of being clinical efficacious while still minimizing any potential disruption to the existing operating room workflow. The following steps include a subsequent Phase 2A proof-of-concept pilot study in patients undergoing PVI under PSA to inform the feasibility and acceptability of the MBI. (ClinicalTrials.gov ID: NCT05837481)

## Data Availability

All data produced in the present study are available upon reasonable request to the authors

## Acknowledgments

We would like to thank Jon Kabat-Zinn, Ph.D., for providing consultation regarding our study and for personally editing a preliminary iteration of our guided meditation script.

